# A thematic analysis of middle managers’ perceptions of a conceptual framework for person-centred outpatient care

**DOI:** 10.1101/2025.10.31.25339226

**Authors:** Fredrik Gasser, Peter Hagell, Albert Westergren, Sidona-Valentina Bala, Lina Behm

## Abstract

**Introduction:** Persons with long-term conditions (LTCs) have been identified as a group that could benefit from more person-centred care (PCC). However, the complexity of PCC as a concept, combined with the scarcity of a shared understanding of its definition across different healthcare contexts, has challenged systematic evaluation. To address this gap, a conceptual framework, operationalised in a patient-reported instrument: the Person-Centred Care instrument for outpatient care (PCCoc), was developed to facilitate evaluation and guide targeted efforts to strengthen PCC in outpatient settings for persons with LTCs. Yet, few studies have explored how middle managers, who play a crucial role in implementing and sustaining organisational development, perceive the clinical applicability of such conceptual frameworks.

**Aim:** To explore middle managers’ perceptions of a conceptual framework for person-centred outpatient care and its clinical applicability.

**Methods:** A qualitative design was applied, using thematic analysis to describe middle managers’ perceptions. Data were collected through semi-structured interviews with middle managers in various outpatient clinics.

**Results:** Three main themes and seven sub-themes were identified: (1) Factors that may facilitate use, (i.e., aspects that may enhance the adoption and application of the conceptual framework); (2) Challenges for practical application, anticipated challenges in applying the conceptual framework in the outpatient setting; and (3) Practical usefulness in a clinical context, perspectives on the potential use of the conceptual framework in outpatient care for persons with long-term conditions.

**Conclusion:** The findings suggest that the conceptual framework is perceived as clinically relevant and may, through its associated patient-reported instrument, PCCoc, facilitate the evaluation and development of person-centred outpatient care. While middle managers recognised its alignment with existing practices and its potential to advance PCC in outpatient setting, challenges to its adoption were also identified, particularly in relation to organisational factors.

## Introduction

Persons with long-term conditions (LTCs), who are often engaged in prolonged outpatient care, have been identified as a group that would benefit from more person-centred care (PCC) (1–4). Potential benefits include improved management of the LTC, enhanced self-esteem maintenance, and increased patient satisfaction (2–3). However, the complexity of PCC as a healthcare concept, combined with the absence of a shared understanding of its definition across various healthcare contexts, has complicated efforts to evaluate PCC systematically (5–6), a challenge that is particularly pronounced for persons with LTCs in outpatient settings (7–8). This is problematic, as the implementation and sustained application of PCC depend on ongoing monitoring and evaluation (9–10), which is essential for ensuring quality and supporting continuous improvement (11).

To address this shortcoming, the Person-Centred Care instrument for outpatient care (PCCoc) was developed (12–13). This patient-reported experience measure (PREM) is intended for persons with LTCs aiming to support the evaluation and advancement of PCC in outpatient settings. The instrument is based on a conceptual framework (12) that focuses on the encounter between the healthcare professional (HCP) and the patient, and characterises person-centred outpatient care along a hierarchical continuum from lower to higher levels of PCC (Fig. 1). As such, it has the potential to facilitate evaluation and targeted efforts to develop PCC in outpatient settings for persons with LTCs.

**Figure 1.**
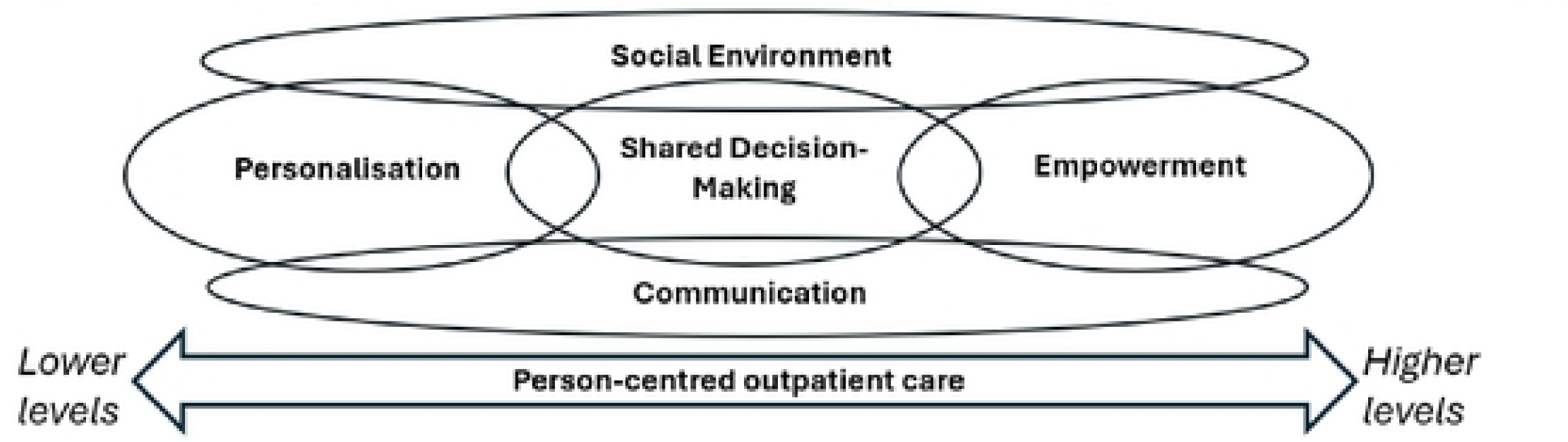
The Conceptual framework for person-centred outpatienl care(reproduced from Gasser et al. (14).

The conceptual framework comprises three overlapping domains that represent a hierarchical continuum, ranging from lower (easier to achieve) to higher (more challenging to achieve) levels of person-centred outpatient care, progressing from personalisation, through shared decision-making, to empowerment (12). Empirical PCCoc data from persons with LTCs in various outpatient settings have been shown to support this hierarchical continuum (14). Social environment and communication are not considered part of the hierarchical continuum, but integrated throughout.

Middle managers hold an important intermediate role between senior management and frontline HCPs, with responsibilities that include implementing evidence-based practices, translating strategic guidelines into clinical improvements, and evaluating both the implementation process and its outcomes (15). In a previous study regarding middle managers’ conceptions of opportunities and barriers of working with PCC in outpatient settings, PCC was conceived to be driven by three interdependent and partially overlapping levels, spanning from the organisational level to the healthcare professional and patient levels, in which the role of middle managers is central (16). Successful implementation and evaluation of person-centred outpatient care are, therefore, arguably dependent on outpatient middle managers’ perceived acceptability and applicability of the underpinning conceptual PCC framework. That is, to facilitate the development of PCC in practice, the conceptual framework must be seen as relevant, understandable, and useful by those implementing and applying its principles in everyday practice and quality improvement. However, little is known about middle managers’ perspectives on the clinical applicability of PCC frameworks, such as that underpinning the PCCoc. This gap limits our understanding of its practical utility and its potential to guide meaningful use and assessment of person-centred outpatient care.

### Aim

To explore middle managers’ perceptions of a conceptual framework for person-centred outpatient care and its clinical applicability.

## Methods

### Design

The study spanned one year and employed a qualitative descriptive design based on interviews. Reflexive thematic analysis was undertaken within a realist (essentialist) orientation and guided by an inductive approach (17) to explore middle managers’ perceptions of a conceptual framework for person-centred outpatient care and its clinical applicability, with data analysed at the semantic level. The consolidated criteria for reporting qualitative research (COREQ) (18) were used in this study.

### Setting

In Sweden, the responsibility for health care is shared between the national government, the 21 regional authorities, and the 290 municipalities, where the regions are primarily responsible for providing, organising, and financing health care. Outpatient care is provided in both primary care and specialist care and involves a range of professional groups. Physicians and nurses are the most common, but other categories, such as physiotherapists, psychologists, nursing assistants, and speech therapists, are also represented (19). Primary care refers to non-hospital care for all patient groups, regardless of age or medical condition. Specialist care covers both somatic and psychiatric conditions and usually requires a referral from a licensed healthcare professional (20). In 2021, there were just over 86 million healthcare encounters in outpatient care in Sweden (19).

A total of 16 different regionally run outpatient clinics were included in this study. The outpatient clinics were located in three geographically dispersed regions in southern and central Sweden, with a total population of approximately 4.23 million, and included both urban and rural areas. The 16 outpatient clinics covered 10 different areas, comprising both primary care and specialisations in somatic and psychiatric outpatient care.

### Participants

This study builds on a previous study among sixteen middle managers, defined in this study as individuals holding formal roles positioned between senior leadership and frontline clinical professionals (21), from various outpatient settings. The recruitment period for this study started in 23/05/2024 and ended 04/28/2025. A strategic sampling was used to ensure variation in both the type of outpatient care (e.g., psychiatric, somatic, and primary care) and geographical distribution. For a more detailed description of recruitment process, see Gasser et al. (16). Following the initial interview (Phase I), all sixteen middle managers were invited to a follow-up individual interview (Phase II), specifically targeting the conceptual framework, in which seven participated (Table 1).

**Table 1.**
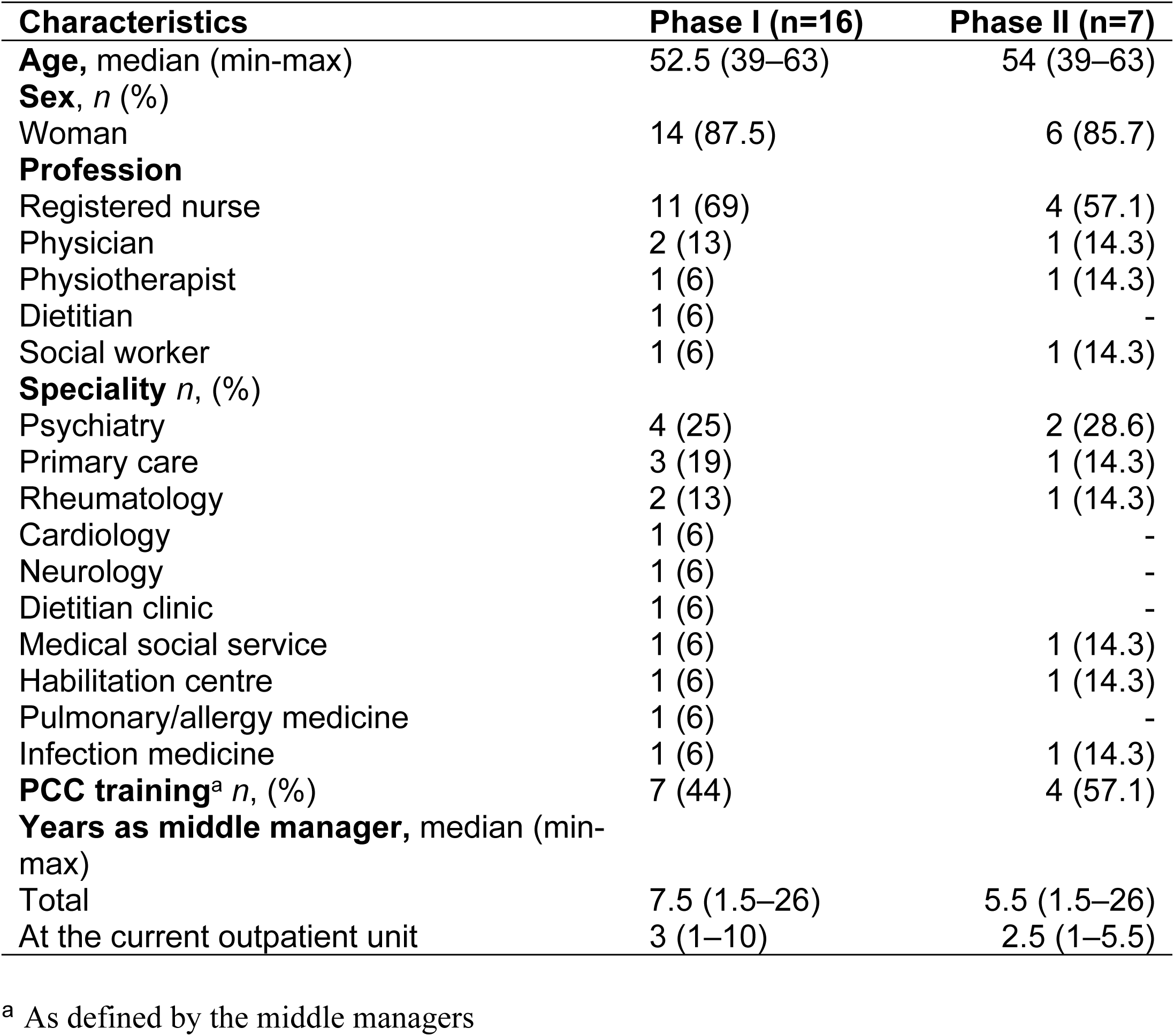
Characteristics of the participants in Phases I and II.

### Data Collection

Data were collected between May 2024 - May 2025 through semi-structured interviews, conducted in two sequential phases. In Phase I (n=16), participants first completed an interview focusing on their perceptions of the opportunities and barriers to working with PCC in outpatient settings (16). Following this, they were presented with a visual representation of the conceptual outpatient PCC framework (Fig. 1), given a brief explanation of its domains, and asked to respond to two questions (Table 2). In Phase II (n=7), participants received the conceptual framework (Fig.1), along with a brief description of its domains (Table 2), via email prior to the interview. During the interview, a study-specific interview guide (Table 2) was used to explore participants’ perceptions of the conceptual framework in greater depth. All interviews were conducted digitally via Microsoft Teams or Zoom by FG or LB, and were recorded and transcribed verbatim. The Phase II interviews lasted between 18 and 40 minutes (mean: 25 minutes).

**Table 2.** Description of the conceptual framework given to the participants and the interview guides.

| Description of the conceptual framework given orally (Phase I) and in writing (Phase II) | Interview guide Phase I | Interview guide Phase II |
| --- | --- | --- |
| The conceptual framework defines person-centred outpatient care from personalisation through shared decision-making to empowerment. Lower levels represent aspects of person-centred outpatient care that are relatively easy to achieve, while higher levels are more difficult to achieve. Personalisation means identifying and recognising the unique needs and abilities of the person. Shared decision-making involves collaboration between healthcare professionals and the person in need of care, where information is shared and healthcare professionals support decision-making. Empowerment is about strengthening the person's own ability to manage and control their situation. Social environment refers to both how a person is treated and the physical environment in which care is provided, and communication means creating a dialogue together that builds a relationship and mutual understanding. | What is your perception of the conceptual framework?<br>Would the possibility to measure the degree of person-centred care from less to more perceived person-centredness contribute anything to your outpatient care unit? | What was your perception of the conceptual framework?<br>How does the conceptual framework correspond to your view of person-centred outpatient care?<br>Is there anything you think is missing from the conceptual framework for person-centred outpatient care?<br>How could the conceptual framework be used in your outpatient care practice?<br>What challenges do you see in implementing this conceptual framework in practice? |

### Data Analysis

Data were analysed inspired by Braun and Clarke’s six-phase process of thematic analysis (17). In phase one (*Familiarisation*), FG and LB immersed themselves in the data by reading all transcripts multiple times and noting initial ideas. In phase two (*Generating initial codes*), descriptive codes were generated systematically by FG through open coding across the dataset. In phase three (*Searching for themes*), codes were organised by FG and LB into potential themes and sub-themes. In phase four (*Reviewing themes*), the initial themes were refined by all authors to ensure internal coherence and that they accurately represented the dataset. In phase five (*Defining and naming themes*), clear and concise names and definitions were developed to reflect the central idea of each theme, and in phase six (*Producing the report*), the final analysis was written up, using illustrative quotes to support each theme.

Although the analysis primarily focused on manifest content, a reflexive approach was acknowledged throughout. The researchers continuously reflected on their role in the analysis process and acknowledged that theme development is to some extend shaped by interpretative decisions (22).

## Results

Three main themes and seven sub-themes were identified from the middle managers’ perceptions of a conceptual framework for person-centred outpatient care and its clinical applicability (Table 3). The three main themes identified were: 1. *Factors that may facilitate use* represents aspects that may enhance the adoption and application of the conceptual framework. 2.*Challenges for practical application* represents anticipated challenges in applying the conceptual framework in the outpatient setting. 3. *Practical usefulness in a clinical context* represents perspectives on the potential use of the conceptual framework in outpatient care for persons with long-term conditions.

**Table 3.**
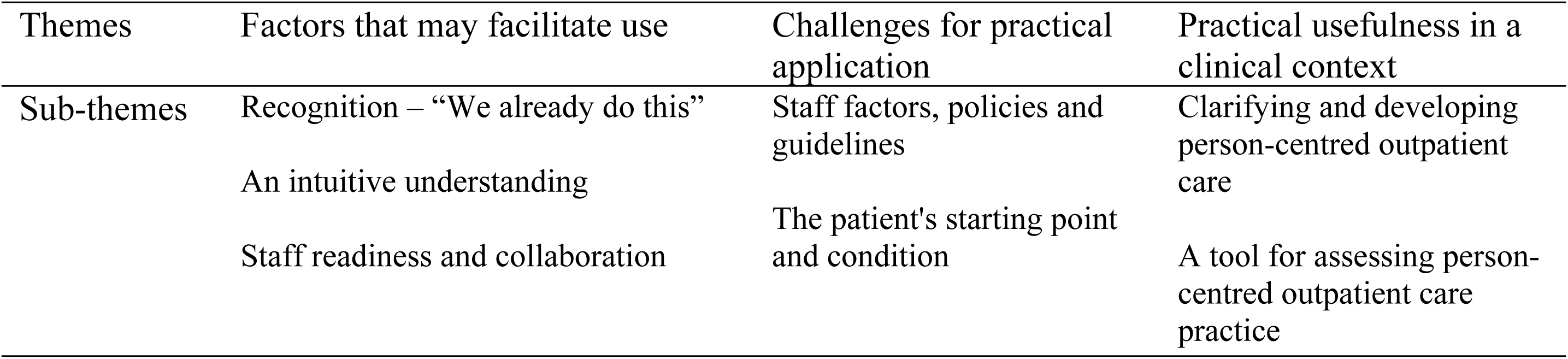
Themes and sub-themes.

### Factors that may facilitate use Recognition – “We already do this”

The middle managers perceive that the conceptual framework and its progression from personalisation via shared decision-making to empowerment is in line with their current approach to PCC. Outpatient care for persons with LTC, with continuous meetings and long-term relationships, is described as particularly conducive to putting the principles of the framework into practice. The middle managers describe how their existing view of the patient, that they ‘own their illness’ and should have power over their own care process, is clarified in the framework.

The framework is also perceived as a reflection of the current way of working, making the organisation’s goals explicit, and is readily recognised as familiar:

“*It doesn’t feel unfamiliar given where we are today… this image confirms what we want to work on.*” Participant 10

The quote illustrates how the conceptual framework functions as a validation mechanism; it confirms that the organisation is already engaged in a process towards PCC and thus anchors continued developments in clinical practice. In the interviews, the middle managers use the conceptual framework as a map, making explicit their current position and the further path required to empower patients, that is, what they need to work on going forward.

### An intuitive understanding

The middle managers consider the conceptual framework to be intuitively understandable. The visualisation of the degree of person-centredness from personalisation to empowerment facilitates understanding, where the “overlap” between the domains clarifies that the domains are interconnected and cannot be viewed as isolated parts. It is also emphasised that the conceptual framework clarifies that PCC entails more than merely providing a respectful encounter. The middle managers express that the fact that PCC is visualised can facilitate implementation or further development of PCC to a greater extent. One middle manager expressed it as follows: “*I believe that we are carrying out highly person-centred work at the clinic. And I consider that the conceptual framework is clear, at least to me, and that it can certainly be used by us as a description, a visual representation of such work*.” Participant 15 Personalisation, which is the first level in the conceptual framework, is perceived as more foundational and easier to achieve, whereas empowerment appears to be more challenging:

> “It does in fact demand more from both healthcare professionals and patients, I think. As healthcare professionals, one needs to step back somewhat and consider the broader context, while also supporting the patient in being able to take that step. Sometimes patients already have this capacity or receive other forms of support that help them, but often it is we who must lift them to that level. So, shared decision-making, absolutely, we are very much engaged in that, and some patients also in empowerment. But for the majority, I would say it probably remains at the level of personalisation.” Participant 14

The middle managers also raise concerns regarding the potential abstraction, ambiguity, or obsolescence of the concepts within the conceptual framework. For example, “social environment” is mentioned as a difficult and ambiguous concept, both for staff and patients. “communication”, “social environment” and “shared decision-making” raised questions about what they actually mean in practice. One manager expressed that it requires reflection to understand the meaning:

> “So what does this mean, communication and social environment and shared decision-making and so on? It requires a bit of thought; it’s not something you understand immediately when you see it.” Participant 9

However, the supporting explanatory text accompanying the framework is perceived as facilitating understanding and thereby the use of the conceptual framework: “*And then it’s really good with these rather short and relatively simple definitions. Because it doesn’t become too pompous or difficult to grasp*.” Participant 14

### Staff readiness and collaboration

The middle managers argue that the introduction of a person-centred framework does not occur in isolation, but is contingent upon staff’s pre-existing knowledge and attitudes. When staff possess a foundational understanding of PCC, such as the importance of patient partnership and shared decision-making, it promotes both comprehension of, and adherence to, the practices required in everyday clinical work. A positive attitude towards PCC also facilitates and makes staff more inclined to try new ways of working, share good examples and support each other when challenges arise.

” *If you consider the concept of person-centred care…it was perhaps something that actually met more resistance in terms of the terminology, maybe around ten years ago. Back then, I remember hearing people getting angry and upset when it was used.*” Participant 11

The middle managers also emphasise that involving the patient is not the responsibility of a single professional, but requires collaboration among multiple professional groups and other services to provide care. They point out that different professional groups contribute distinct perspectives and that no single profession can address all of a patient’s needs independently. Interprofessional collaboration is therefore essential to facilitate the implementation of PCC and to support its ongoing development.

“*We think more broadly in a way, that it is not me, but us who should help our patients to feel better.*” Participant 3

### Challenges for practical application Staff factors, policies and guidelines

The middle managers highlight that the usefulness of the conceptual framework is largely influenced by both staff conditions and the organisational context. For example, it was perceived as challenging to meet patients’ requests when the organisation’s capacity is limited: “*It is far from everything we can fulfil. There are frameworks, boundaries, and limitations to what we can do, so patients are not always guaranteed to be satisfied with the decisions that are made*” Participant 8

A recurring perception is that staff from different professional categories have varying opportunities to provide care with a high degree of person-centredness. For example, it was perceived that certain professions adopt a more natural person-centred approach, whereas in others, more administrative or medical aspects predominate. One middle manager expressed it as follows: “*Well, I think that if you go to a psychologist, it might be very much about empowerment. If you go to a physician, it’s more like, yes, we’ll arrange sick leave*.” Participant 3

Another key aspect was time, both in terms of the time required to understand what the patient actually wants and needs, and the fact that delivering care with higher degrees of person-centredness necessitates continuity and opportunities for reflection. Several middle managers felt that the organisation’s current structure and resource allocation often do not allow for this in practice. “*Because you don’t really feel you have time for anything other than meeting with patients. So, it’s not easy to have these more philosophical and theoretical discussions*.” Participant 14

Lack of time was also identified as an obstacle to both communication and the ability to create the unique encounters on which PCC is based. One participant expressed this as follows: “*And this thing about how we communicate, where lack of time can sometimes affect it. But we need to remember that every conversation is unique.*” Participant 12

The physical care environment is described as an important component of PCC, yet at the same time challenging to modify, even when there is an intention to improve it. For example, this is influenced by the available space in existing facilities, hygiene regulations, and financial constraints, as the budget for implementing changes in the care environment is limited.

The middle managers emphasised that applying the conceptual framework requires a clear purpose for how it should be used and understood within each outpatient unit. This may necessitate training initiatives, various implementation strategies such as workshops and informational materials, as well as long-term objectives, as initiating and sustaining changes in operations is not a quick fix.

”*I think that if you’re going to implement it, you need to be very clear about what it’s for, that it’s very clearly stated. Because I think anyone in my outpatient unit who had looked at this would have said: yes, absolutely, and then what?*” Participant 14

### The patient’s starting point and condition affect

The middle manager emphasises that outpatient care can almost always be personalised, but the patient’s clinical condition, socio-economic situation, and level of education can influence the possibility of achieving shared decision-making and empowerment.

“*It is very clear how much the patient’s basic condition and functioning, both mentally and in terms of frailty, play a role. You may want the person to have as much influence as possible, but certain factors sometimes limit this possibility*” Participant 1

They also point out that some patients prefer more directed care, which requires that care be adapted to the patient’s individual capacity and preferences. For example, one middle manager highlights that several patients express the opinion:

> “ I want you to make the decisions based on what you already know about me and my situation. I leave it to you; I don’t want to be involved.” Participant 11

Some middle managers believe that the conceptual framework is easier to apply to patients with complex conditions, where it is clearer how to work towards higher levels of person-centred outpatient care.

” *I think that the patient you have in front of you will determine how far we get. The more complex the patient’s needs are that can be supported, the easier it will be for us to achieve a higher level.*” Participant 10

Another tendency that has been noted is an emphasis on patients managing their own situation. However, such an emphasis may sometimes be problematic, as healthcare providers need to assume greater responsibility when patients are not considered fully able to do so independently.

### Practical usefulness in a clinical context Clarifying and developing person-centred outpatient care

The middle managers perceive that PCC can be difficult to implement in clinical work, and there may be varying perceptions among staff as to what it means. They emphasise that the conceptual framework can be a tool for visualising what person-centred outpatient care meetings can entail and strive for, but also for highlighting aspects of PCC that already exist in the organisation, which currently is lacking.

”*I would use the conceptual framework to highlight that we are on the right track. That the things we do that we may not think about are actually person-centred outpatient care. But we have never really had any illustration of that*.” Participant 15

By initiating training initiatives and staff discussions based on the conceptual framework, there is an opportunity to establish a common terminology and a shared understanding of how person-centred outpatient care can be realised and further developed in clinical practice. For example, through discussions about what shared decision-making and empowerment mean, both more broadly within the outpatient care unit and for specific patient groups. Such discussions also provide an opportunity to identify the efforts required to promote person-centred outpatient care and enhance patient participation, as reflected in the following quote:

> ”The conceptual framework had been helpful for discussing, okay, we have this group of patients with these abilities. What does empowerment mean for them, and how might we reach that point? How can we facilitate the process?” Paricipant 14

It is also emphasised that the conceptual framework can be used in patient consultations and information brochures to clarify what person-centred outpatient care can entail, but also as a basis in individual patient consultations to initiate discussions on how the patient and healthcare professional can co-design the care.

### A tool for assessing person-centred outpatient care practice

The middle managers stress the importance of evaluating PCC in order to improve practice. The conceptual framework is considered a useful tool for assessing strengths, identifying areas for improvement, and monitoring both improvement efforts and overall objectives, with the aim of promoting person-centred outpatient care. As one middle manager put it: ”*It’s kind of a confirmation of how we work… where do we stand (in relation to the conceptual framework), do we need to improve, or are we working as well as we possibly can given the circumstances*” Participant 6

The fact that the conceptual framework, with its hierarchical structure ranging from lower to higher levels of PCC, can serve as a common foundation for assessment is considered a strength. The middle managers emphasise the importance of highlighting patients’ experiences and perceptions, for instance to identify any discrepancies between staff perceptions and patients’ experiences of person-centred care.

”*That’s how we find out, if we think something, we get it in black and white when we have questionnaires where the patients themselves can report on their experience*” Participant 3

At the same time, the middle managers highlighted that the hierarchy in the conceptual framework, with its different levels and scale, can create uncertainty, particularly regarding what is considered “higher” or “lower” in clinical practice. One suggestion was to have the arrow in the conceptual framework point in only one direction and closer to the “three circles” representing the levels of personalisation, shared decision-making, and empowerment. There were also varying interpretations of the hierarchy. For example, some questioned why “empowerment” should be placed higher than “shared decision-making”, while others argued that empowerment should rather permeate all parts of the framework instead of being placed as an end goal. They also emphasised the relational aspect as a prerequisite for achieving shared decision-making and empowerment, noting that it is not explicitly highlighted in the conceptual framework:

> ”I think the relational aspect is really part of both communication and personalisation; it’s kind of a prerequisite for all of these things, really. But maybe it’s so obvious that it’s not something that needs to be highlighted on its own, and you just have to think: well, it’s a prerequisite for all of this to work.” Participant 11

## Discussion

Leadership has been highlighted as a key component in the implementation and development of PCC (23–24), with middle managers recognised as having an important role (15). However, there appears to be a lack of studies investigating how middle managers perceive the conceptual frameworks that often underpin the conceptualisation of PCC in the intended context. We therefore explored middle managers’ perceptions of a conceptual framework for person-centred outpatient care and its clinical applicability, through which three themes were identified: Factors that may facilitate use; Challenges for practical application; Practical usefulness in a clinical context.

### Factors that may facilitate use

The conceptual framework for person-centred outpatient care was seen as aligning with the middle managers’ perspectives on PCC in an outpatient context and their current working practices, which may facilitate its use and potential implementation. PCC can be perceived as an abstract concept that is difficult to define (25–27) and the importance of being able to operationalise abstract principles in clinical practice has been highlighted in the implementation of PCC (28). The perceived clarity of the framework among the middle managers can serve as a facilitating factor in understanding what person-centred encounters in outpatient care may entail for persons with LTCs. It may therefore act as a bridge between the abstract nature of PCC and how it can be operationalised in everyday practice.

An important finding was that the conceptual framework was perceived as adapted and useful for the intended target group, persons with LTCs. Williams et al. (29) emphasise that PCC for this group must be tailored to the challenges posed by managing a condition over an extended period, which often has a significant impact on daily life. Self-management of the condition is recognised as an important component, which requires that the person has knowledge about the condition and receives support from healthcare services for empowerment. At the same time, there are signs that persons with LTCs experience healthcare as being less person-centred, for example, due to insufficient involvement in their care (30). The conceptual framework may therefore serve as a useful foundation for developing interventions that strengthen PCC for persons with LTCs.

Although middle managers perceived the conceptual framework to be in line with their view of PCC, certain concepts within it, such as shared decision-making and communication, were perceived as complex. This is not surprising since conceptual frameworks often operate at a higher level of abstraction (31). This implies that the organisation needs to concretise what the conceptual framework may entail in the specific context. At the same time, the discussions it initiates may constitute an important component in creating a shared understanding and conceptual language for PCC in that context, which can be seen as a prerequisite for implementation (28; 32).

### Challenges for practical application

The middle managers perceived potential challenges in applying the conceptual framework in clinical practice. These challenges related broadly to organisational factors and resources, including staff conditions, inter-professional collaboration, and organisational priorities that allow time and space for person-centred conversations. The importance of organisational factors has been addressed in Santana’s framework for implementing PCC, which identifies a work environment with sufficient resources as a prerequisite (32). Similarly, McCormack et al. emphasise that a person-centred culture permeating the entire organisation is essential for strengthening person-centred processes (33). McCormack also critiques evaluations of person-centred care that fail to consider the prevailing organisational culture (34). Although the conceptual framework does not explicitly adopt an organisational perspective, it can shed light on practical challenges, acting as a tool to advance person-centred outpatient care for persons with long-term conditions (12).

The patient’s starting point and medical condition were also perceived as potential challenges. These were primarily related to the domains defined as higher levels of person-centred outpatient care, such as shared decision-making and empowerment. For example, it was suggested that patients’ socio-economic status and level of education might influence the feasibility of achieving these higher levels of person-centred care within the conceptual framework. At the same time, PCC has been shown to enhance participation among individuals with lower levels of education (35), highlighting the need for ongoing development and evaluation of PCC in outpatient settings to better understand and map its possibilities.

Middle managers also expressed concerns that a conceptual framework progressing from lower to higher levels might create pressure to reach higher levels even when this is not feasible or desirable. While this is a valid and important point, it is crucial that efforts to promote aspects such as shared decision-making are not compromised due to perceived difficulties associated with specific patient groups (36).

### Practical usefulness in a clinical context

A key finding was that middle managers perceived the conceptual framework as a helpful tool for clarifying, developing and assessing PCC for persons with LTCs in outpatient care. This is significant because the operationalisation of the conceptual framework in a PREM (the PCCoc) (12–13) corresponds with the hierarchy in the conceptual framework, from lower to higher levels of PCC (14). This enables ongoing assessments from the patients’ perspective, which can facilitate targeted interventions addressing challenges within a specific domain (personalisation, shared decision-making, or empowerment) to strengthen the domain and further develop person-centred outpatient care. Furthermore, assessing care from the patient’s perspective has been highlighted as valuable for identifying care-related issues, and supporting staff reflection at individual, team, and organisational levels. It can also guide service restructuring while monitoring the impact of changes (11), all of which have implications at an organisational level. Accordingly, the conceptual framework and PCCoc can serve as a tool in the internationally growing intention to ensure person-centred healthcare (9). An intention that has been hampered by a lack of consensus on the most suitable instruments (5), as well as the absence of a clear theory or conceptual framework underpinning the instruments, resulting in ambiguity regarding what is actually being assessed (34). Although instruments have been developed to evaluate person-centred care based on frameworks (e.g. 37), the underlying frameworks rarely adopt a hierarchical structure from lower to higher levels, which can complicate clinical application due to the absence of guidance and prioritisation of potential actions. Furthermore, without an idea of the nature of progression from less to more, the interpretation and meaning of results from the application of such tools remain ambiguous.

### Limitations

Outpatient care is multifaceted, and our selection of outpatient care units is open to question. At the same time, we have included medical and psychiatric specialist care as well as primary care in order to obtain as good a representation of outpatient care as possible. Of the 16 middle managers who participated in the first phase, nine chose not to participate in the follow-up interview, meaning that we do not have in-depth knowledge of their perceptions of the conceptual framework.

Braun & Clarke have highlighted that interpretations are inevitable and part of the analysis (22). All authors have clinical experience of outpatient care and, with the exception of LB, are well acquainted with the conceptual framework. LB’s prominent role in the analysis process can therefore be seen as a strength in that it balances preconceptions. During the analysis process, the authors reflected on the assumptions underlying the analysis, i.e. a realist (essentialist), inductive and semantic approach, with an endeavour to stay close to the respondents’ own perceptions, well aware that this narrows the scope of the reflexive thematic analysis (38).

## Conclusion

Research examining how middle managers perceive conceptual frameworks for PCC is scarce, a limitation given their pivotal role in implementing and sustaining organisational change. The present study explored middle managers’ perceptions of a conceptual framework for person-centred outpatient care. The findings indicate that the framework was perceived as being aligned with the middle managers’ views on person-centred outpatient care and their existing working practices, and as a potentially valuable tool for clarifying and advancing person-centred practices within outpatient care settings. This supports the clinical relevance of the framework and that its associated PREM (the PCCoc) may facilitate the evaluation and development of person-centred outpatient care. Nevertheless, challenges to its adoption were identified, predominantly related to organisational factors, but also to the conceptual framework.

**Declarations**

## Ethics approval

The study was approved by the Swedish Ethical Review Authority (Dnr. 2021-00620; approval date February 22, 2021; supplementary application (Dnr 2024-02981-02) approved June 23, 2024. The study was conducted in accordance with the Declaration of Helsinki.

## Consent to participate

Written informed consent was obtained from all study participants.

## Data availability

The datasets used and analysed during the current study are available from the corresponding author on reasonable request.

## Author contributions

All authors contributed to the conception and design of the study. Data collection and initial analysis were carried out by FG and LB and subsequently reviewed and refined in consultation with the other authors. The manuscript was drafted by FG, with all authors providing feedback during its development.

## Acknowledgements

The authors want to thank the participating middle managers for their time and collaboration. The study was supported by Kristianstad university, Kristianstad, Sweden.

